# The Impact of Early or Late Lockdowns on the Spread of COVID-19 in US Counties

**DOI:** 10.1101/2021.03.19.21253997

**Authors:** Xiaolin Huang, Xiaojian Shao, Li Xing, Yushan Hu, Don D. Sin, Xuekui Zhang

## Abstract

**Background:** COVID-19 is a highly transmissible infectious disease that has infected over 122 million individuals worldwide. To combat this pandemic, governments around the world have imposed lockdowns. However, the impact of these lockdowns on the rates of COVID-19 transmission in communities is not well known. Here, we used COVID-19 case counts from 3,000+ counties in the United States (US) to determine the relationship between lockdown as well as other county factors and the rate of COVID-19 spread in these communities.

**Methods:** We merged county-specific COVID-19 case counts with US census data and the date of lockdown for each of the counties. We then applied a Functional Principal Component (FPC) analysis on this dataset to generate scores that described the trajectory of COVID-19 spread across the counties. We used machine learning methods to identify important factors in the county including the date of lockdown that significantly influenced the FPC scores.

**Findings:** We found that the first FPC score accounted for up to 92.81% of the variations in the absolute rates of COVID-19 as well as the topology of COVID-19 spread over time at a county level. The relation between incidence of COVID-19 and time at a county level demonstrated a hockey-stick appearance with an inflection point approximately 7 days prior to the county reporting at least 5 new cases of COVID-19; beyond this inflection point, there was an exponential increase in incidence. Among the risk factors, lockdown and total population were the two most significant features of the county that influenced the rate of COVID-19 infection, while the median family income, median age and within-county move also substantially affect COVID spread.

**Interpretation:** Lockdowns are an effective way of controlling the COVID-19 spread in communities. However, significant delays in lockdown cause a dramatic increase in the case counts. Thus, the timing of the lockdown relative to the case count is an important consideration in controlling the pandemic in communities.

**Research in context:** *Evidence before this study:* We searched PubMed using the term “coronavirus”, OR “COVID-19”, OR “COVID-19 infection”, OR “SARS-CoV-2” combined with “Lockdown” or “sociodemographic factor” or “Vulnerability” for original articles published before March 18, 2021. Similar searches were done in medRxiv, Google Scholar, and Web of Science. Only papers published in English were reviewed. The most similar relevant works to our study were Acharya et al.^1^ and Karmakar et al.^2^, which investigated the associations between population-level social factors and COVID-19 incidence and mortality. Unlike our current study, which employed a longitudinal design, both of studies were cross-sectional in nature and thus fixed on a single time point. In addition, neither of these studies investigated the impact of lockdown measures on COVID-19 infection patterns. Another relevant study is Alfano et al.’s work3, which focused on the efficacy of lockdown on COVID-19 case rates. However, this study did not evaluate the timing of lockdown on this endpoint.

*Added value of this study:* To our knowledge, this is the first study to use functional principal component analysis (FPCA) to investigate COVID-19 infection trajectories (in a longitudinal manner) and their relationships with different sociodemographic factors and lockdown policy at a county level. The FPCA transformed a longitudinal vector with high-dimensions into a “single” surrogate variable, which retained 93% of the information. We used an advanced statistical model (segmented regression) to investigate the effects of lockdown on incidence of COVID-19 across the US. We found that the relationship had a “hockey stick” appearance with an inflection point at ∼7 days prior to a county reporting at least 5 cases of COVID-19. We also applied a machine learning model (i.e., elastic net) to explore joint effects of lockdown and other sociodemographic factors on COVID-19 infection patterns, which estimated the impact of each of factors, adjusted for each other.

*Implications of all the available evidence:* Our study suggests that lockdown is an effective policy to reduce case counts of COVID-19 in communities; however, significant delays in its implementation result in exponential growth of COVID-19. The inflection point is approximately 7 days prior to a county reporting at least 5 cases of COVID-19. These data will help policy-makers to determine the optimal timing of lockdowns for their communities.

## INTRODUCTION

Coronavirus 2019 (COVID-19) is a global pandemic that has affected over 122 million individuals and killed 2.7 million people across the world^4^. Compared with the original severe acute respiratory syndrome (SARS), the SARS-CoV-2, the virus responsible for COVID-19, is much more contagious with a basic reproductive ratio (commonly denoted as R0) of 2-4^5^ in most countries. At this R0, there is an exponential growth in the case counts of COVID-19 in the community, leading to large increases in COVID-19 related morbidity and mortality, which may overwhelm local health care systems. To address this issue and to reduce the spread of COVID-19, governments around the world have imposed lockdowns (at a varying intensity) on their communities^6^ with some imposing lockdowns early in the pandemic, while others much later. However, the impact of the timing of the lockdown relative to the first few reported cases of COVID-19 in communities is largely a mystery. Beyond lockdowns, it is not well-known what features of communities significantly influence the rate of COVID-19 spread across jurisdictions. Here, we used data from counties in the United States (US) to determine the impact of lockdown timing on COVID-19 spread and identify potential features of communities that influence the rate of COVID-19 spread across these counties.

## METHODS

### COVID-19 case counts during the pandemic across the United States (US) counties

We extracted COVID-19 data from the Johns Hopkins Coronavirus Resource Center^7^ and analyzed the daily records of cumulative COVID-19 case counts across 3,340 counties in the United States (US) from 2020-01-22 to 2021-01-31. We excluded counties that were not included in the US American Community Survey (ACS)^8^ 5-year estimates, leaving 3,140 counties in the dataset. We further excluded counties that did not report at least five total cases of COVID-19. Thus, the final dataset contained data from 3,116 counties. We realigned the dataset to ensure that there were at least five cases per county at the start of observation time. We denoted Q_*ij*_ as the infection count in the *i*^*th*^ county on the *j*^*th*^day.

### Modeling the spread of COVID-19 over time in the US counties using unsupervised machine learning

To investigate the pattern of infection spread over time in each of the counties, we applied a functional principal component (FPC) analysis^9^. The FPC model is given as the following formula:

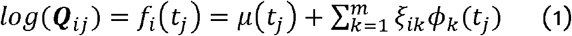

We considered the observed cumulative counts Q_*ij*_ for the *i*^*th*^ county as a trajectory generated from the function *f*_*i*_(*t*_*j*_) and *t*_*j*_ is the *j*^*th*^day, which grew exponentially with varying rates on different days. The FPC model maps these trajectories onto a m-dimensional functional space spanned by orthogonal eigen-functions *ϕ*_*k*_(). Each eigen-function describes how individual trajectories differ from *μ*(), which represents the average trajectory across all US counties. The eigen-functions are ordered by the proportion of variance in the data that can be explained by these functions. The first eigen-function explains the greatest amount of the variance in the dataset; the second eigen-function explains the greatest amount of variance in the dataset that cannot be explained by the first eigen-function; the third eigen-function explains the greatest amount of variance in the dataset that cannot be explained by the first two eigen-functions; and so on. The coefficient *ξ*_*ik*_ is the functional principal component (FPC) score or the coordinate of the *i*^*th*^ county in the *k*^*th*^ dimension of functional space. This score describes the strength of the *k*^*th*^ pattern in the *i*^*th*^ county. Muller et al^9,10^ introduced the theoretical details that outline the method by which estimate functions *μ*() and *ϕ*_*k*_()as well as coefficients *ξ*_*ik*_ are generated. In this work, we estimated these parameters using the R package fdaPACE^11^.

The FPC analysis is a typical unsupervised machine learning approach for dimensional reduction, which reduces high-dimensional data of raw counts Q_*ij*_ (vectors of 376 days) into m FPC scores *ξ*_*ik*_. In the present study, we chose m = 1, which explained 92.81% of the total variance in the data. Thus, we used a single FPC score, *ξ*_*i*1_, to describe the spread of COVID-19 of the *i*^*th*^ county. The larger the value for *ξ*_*i*1_the greater the number of cases of COVID-19. As the rate of COVID-19 spread was not constant over time, we used eigen-function *ϕ*_l_() to describe the rate of the increase. To investigate the impact of various societal factors on the spread of COVID-19 infection in the community, we examined the relationship of these demographics and policy factors with the 1^st^ FPC scores.

### Demographic characteristics of each county and the dates of lockdown according to US counties

We extracted the demographic, socioeconomic, and health data for each county from the 2018 US Census (using R package tidycensus^12^). Specifically, we fetched the following parameters from the American Community Survey (ACS) five-year data profile (see supplementary Table S1) : demographics (comprising total population, male population, population density, and number of families), socioeconomic status (comprising median family income, aggregate family income across the entire county, and the Gini Index), household composition (median age), visible minority status and language (which consisted of race variables), geographical mobility and mode of transportation, and health insurance status (private versus public coverage of health insurance). In addition, for each county, we determined the date on which a lockdown was implemented and calculated the difference between the lockdown date (defined as the date on which a stay-at-home policy was implemented^13^) and the first day in which the county reported 5 or more active COVID-19 cases. To investigate the relationship of these county-level characteristics with COVID spread, we employed a simple linear regression model to test the marginal association between each county-level characteristic (predictor) and the first FPC scores (outcome). We present these results in Table 2.

**Table 1:** Baseline characteristics of counties according to implementation of early or late lockdown (defined as whether or not implementation date is before inflection point, i.e., 7 days before 5 total cases reported in a county) in the course of the pandemic. P-values for all variables are smaller than 2*10^-3^ in Wilcoxon test for difference between early lockdown and late lockdown. Data are shown as mean±SD

| **County Characteristics** | **Early Lockdown**<br>**(n=1346)** | **Late Lockdown**<br>**(n=1385)** |
| --- | --- | --- |
| Total Population ($\times 10^3$) | 20.8±21.5 | 206±476 |
| Population Density (number of people per sq mile) | 64.1±319 | 539±2660 |
| Median Age (years) | 42.7±5.52 | 39.7±4.68 |
| Number of Families ($\times 10^3$) | 5.36±5.45 | 49.7±109 |
| Median Family Income (\$ $\times 10^3$) | 59.5±12 | 67.8±18.4 |
| Aggregate Income of All Families (\$ $\times 10^8$) | 4.02±4.58 | 51±121 |
| Gini Index | 0.441±0.037 | 0.452±0.0343 |
| Proportion of Males in the county (%) | 50.7±2.82 | 49.5±1.85 |
| Proportion of Whites in the county (%) | 87.5±14.4 | 77.3±17.3 |
| Proportion of African Americans in the county (%) | 5.17±11.1 | 14.4±16.8 |
| Proportion of Natives in the county (%) | 2.19±7.61 | 1.09±4.42 |
| Proportion of Asians in the county (%) | 0.739±1.79 | 2.16±3.58 |
| Proportion of Pacific Natives in the county (%) | 0.0714±0.192 | 0.109±0.588 |
| Proportion of Other Races in the county (%) | 2.07±4.71 | 2.31±3.28 |
| Proportion of Individuals who used Public Transport (%) | 0.44±1.03 | 1.54±4.43 |
| Proportion of Individuals who used Bus (%) | 0.376±0.808 | 1.01±1.98 |
| Proportion of Individuals who used Subway (%) | 0.0228±0.474 | 0.318±2.78 |
| Proportion who Moved within the Same County (%) | 5.74±2.47 | 7.14±2.65 |
| Proportion who Moved in from a Different County (%) | 4.5±2.54 | 4.06±2.06 |
| Proportion who Moved in from a Different State (%) | 2.05±1.89 | 2.28±1.61 |
| Proportion who Moved from Abroad (%) | 0.282±0.641 | 0.423±0.511 |
| Proportion with Private Health Insurance (%) | 63.3±9.98 | 66.2±9.9 |
| Proportion with Public Health Insurance (%) | 41.5±8.91 | 37±8.01 |

**Table 2:** The unadjusted relationship of the baseline characteristics of the counties with the COVID-19 spread in these communities. Variables are sorted by R^2^. The first FPC score is used as a surrogate for COVID-19 spread across the counties. (*Results from segmented regression model; the rest are from linear regression models)

| County Characteristics | Association with the First FPC Score |  |  |
| --- | --- | --- | --- |
| | Coefficient | P-value | $R^2$ |
| Lockdown Slope Before the Inflection Point | 0.05239 | 2.38E-09 | 0.4467 * |
| Lockdown Slope After the Inflection Point | 1.9496 | < 2e-16 |  |
| Number of Families | 0.59686 | 5.81E-300 | 0.35593 |
| Aggregate Income of All Families (\$) | 0.58011 | 1.45E-279 | 0.33622 |
| Total Population | 0.57999 | 1.29E-279 | 0.33618 |
| Median Age (years) | -0.4277 | 8.23E-139 | 0.18266 |
| Proportion of Asians | 0.42232 | 4.91E-135 | 0.17809 |
| Proportion who Moved within the Same County | 0.38517 | 1.12E-110 | 0.14804 |
| Proportion of Individuals who Used Bus | 0.36051 | 3.27E-96 | 0.12965 |
| Proportion of Individuals who Used Public Transport | 0.35687 | 3.51E-94 | 0.12704 |
| Proportion of Whites | -0.31254 | 1.43E-71 | 0.09739 |
| Median Family Income (\$) | 0.29529 | 1.07E-63 | 0.086879 |
| Population Density (number of people per square mile) | 0.28964 | 2.78E-61 | 0.083596 |
| Proportion with Public Health Insurance | -0.25049 | 8.68E-46 | 0.062442 |
| Proportion of African Americans | 0.22642 | 1.63E-37 | 0.050962 |
| Proportion of Other Races | 0.22392 | 1.04E-36 | 0.049834 |
| Proportion of Individuals who Used Subway | 0.20512 | 6.21E-31 | 0.041753 |
| Gini Index | 0.20122 | 8.28E-30 | 0.040168 |
| Proportion who Moved from Abroad | 0.19764 | 8.53E-29 | 0.03874 |
| Proportion of Male | -0.17295 | 2.38E-22 | 0.029602 |
| Proportion who Moved in from a Different County | -0.10542 | 3.70E-09 | 0.010793 |
| Proportion with Private Health Insurance | 0.092981 | 2.00E-07 | 0.0083272 |
| Proportion of Natives | -0.052798 | 0.0031973 | 0.0024674 |
| Proportion who Moved in from a Different State | 0.050974 | 0.0044378 | 0.0022771 |
| Proportion of Pacific Natives | 0.032769 | 0.067407 | 7.53E-04 |

### Modeling lockdown effect using segmented regression

The observed relationship between the FPC scores and lockdown policy was non-linear: it was a “hockey stick” shape with an inflection point that denoted a significant change in the slope. Thus, we used segmented regression to model this relationship and to ascertain the inflection point for the lockdown variable. We defined *T*_*i*_ as the difference in days between the date on which the county experienced at least five cumulative cases of COVID-19 and the date on which the county initiated a lockdown. We denote *T*_*i*_ =*β* as the inflection point, where *β* is an unknown model parameter to be estimated. We derived the following three new variables from the lockdown information:

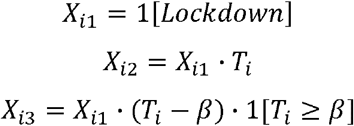

and defined the segmented model as:

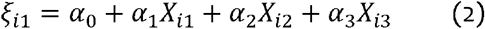

where *X*_*i*1_ (*Lockdown Indicator*) indicated whether a lockdown was implemented for the *i*^*th*^ county, which enabled the model to incorporate data from counties without a lockdown. *X*_*i*2_ (*Lockdown Time*) was the same as the lockdown time *T*_*i*_ when lockdown was implemented by the *i*^*th*^ county; otherwise, it was assigned a value of 0. *X*_*i*3_ (*Lockdown Time after Inflection point*) indicated the date of the lockdown of the *i*^*th*^ county relative to the inflection point. The model (2) is equivalent to three models under different conditions as below:

When a lockdown was not implemented in a county, i.e., *X*_*i*1_ = 0, model (2) simplified into a constant *ξ*_*i*1_= *α*_0_ where the first FPC score is modeled as a constant that is not related to the lockdown time.

When a lockdown was implemented in a county before the inflection point, time *β*, i.e., *X*_*i*l_ = 1 and *T*_*i*_ < *β*, then model (2) simplified into a linear model of *T*_*i*_

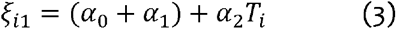

When a lockdown was implemented in a county after the inflection point, time *β*, i.e., *X*_*i*l_ = 1 and *T*_*i*_ > *β*, then the model (2) simplified into another linear model of *T*_*i*_ with a different slope and intercept

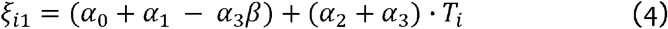

Note, when a lockdown was implemented exactly at the inflection point in a county, i.e., *X*_*i*l_ = 1 and *T*_*i*_ = *β*, then models (3) and (4) become the same model, which ensures that the different slopes in the segmented regression models are well connected at the inflection point. Based on previously discussed models, the coefficients can be explained in the following. The *α*2 shows the effect of a lockdown when it is implemented prior to the county experiencing at least 5 cumulative cases, and *α*_2_ + *α*_3_ represents the effect of the timing of the lockdown when lockdown is implemented after the inflection point. The values of model parameters (*α*_0_, *α*_l_, *α*_2_, *α*_3_, *β*) can be estimated using a profile likelihood approach.

### Modeling joint effect using supervised machine learning

Finally, to explore the joint effects of all risk factors on the first FPC scores, we fitted an elastic net model^14^ to these data. Elastic net is a popular machine learning method, which is based on a regularized linear model.

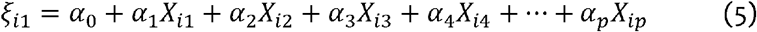

where (*X*_*i*l_, *X*_*i*2_, *X*_*i*3_) are derived variables from lockdown information as defined in model (2), (*X*_*i*4_,…, *X*_*i*p_) are (*p* - 3)demographic characteristics of the *i*^*th*^ county. Elastic net selects important predictors in a linear model (5) by automatically assigning a zero coefficient to unimportant predictors via a penalty on absolute values of coefficients. This penalty also addresses multiple-collinearity among predictors. To capture the uncertainty of the risk estimates, we generated 95% confidence intervals for each coefficient using a re-sample (bootstrap) approach. Specifically, we sampled the counties for replication 1000 times. Next, we applied an elastic net model to each of these random subsets to generate 1000 sets of estimated coefficients, and then built a 95% confidence interval using these coefficients. Here, we fitted all elastic net models using R Package ‘glmnet’^15^. Statistical significance was defined by p-value < 0.05. All data analyses were performed using R Statistical Software^16^ (version 4.0.3). The source codes are available to public by accessing https://github.com/ubcxzhang/COVID.FPCA/

### Interpretation of fitted models

We used two-step modelling, which consisted of (1) unsupervised machine learning (FPC analysis) for dimensional reduction and derivation of surrogate variables, and (2) supervised machine learning (elastic net) to determine the association between risk factors and the FPC scores. When m=1, and by combining these two models, we obtain:

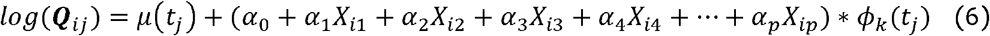

For the *n-th* predictor *X*_*in*_ in model (6), if its value increases by *S*, after adjustment of all other variables, the FPC score increases by *α*_*n*_*S*. This leads to a COVID-19 count, Q, in formula (1), multiplied by exp (*α*_n_*S ϕ*_l_(*t*_*j*_)) in the j-th day after the county reports at least 5 cumulative cases.

## RESULTS

### Functional Principal Component Analysis of COVID-19 Case Counts

We performed a Functional Principal Component (FPC) Analysis on the trajectories of COVID-19 spread across 3,116 US counties. Strikingly, the first FPC explained a vast majority of the total variance (about 92.81%). Figure 1 shows the average trajectory of COVID-19 infection counts across all US counties, which is denoted by the function *μ*(). A positive first FPC score indicates a trajectory that is higher than the national average at any time point; thus, the first FPC score can be considered as a weighted average of infection counts in a county. The varying weights on different days represent the differences in the growth rate of COVID-19 counts in counties across different time points. Figure 2 shows a heat map of the US according to the FPC score for each county. It denotes a time-adjusted COVID-19 case count for each county. The darker colored regions experienced a greater rate of COVID-19 infection. On average, counties, which were in the western and eastern coastal states, had significantly higher rates of COVID-19 infection than those in the central states. The most severely affected counties were found in New York, Arizona, Florida, and California.

**Figure 1:**
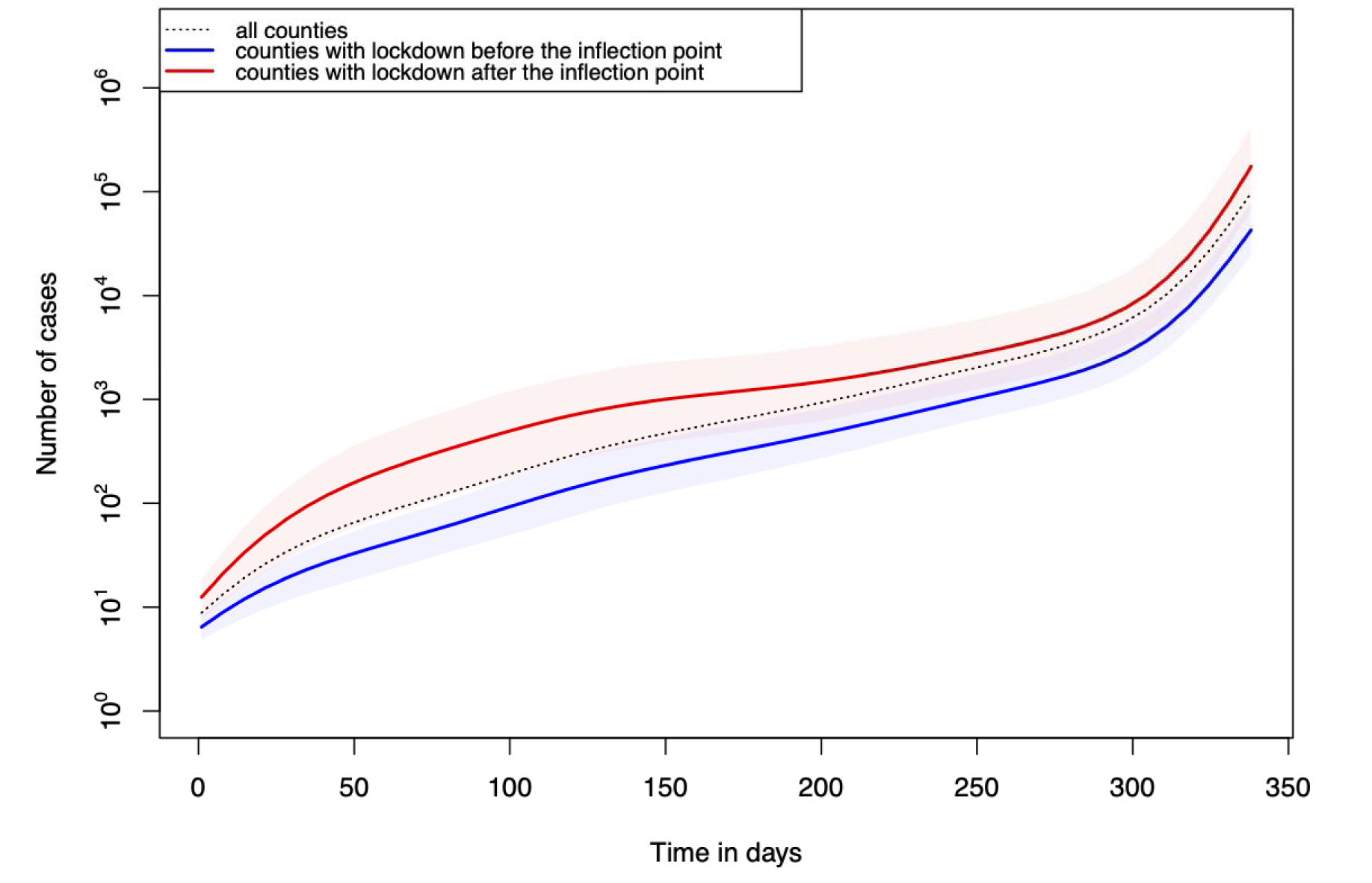
The mean curve of COVID-19 incidence trajectories. The red curve represents the national average of infection cases over time. The green curve represents the average rate of infections of counties that implemented a lockdown before the inflection point. The blue curve represents the average rate of infections of counties that implemented a lockdown after the inflection point. The shaded area represents confidence bound constructed using interquartile range (i.e. 25% −75% quantiles).

**Figure 2:**
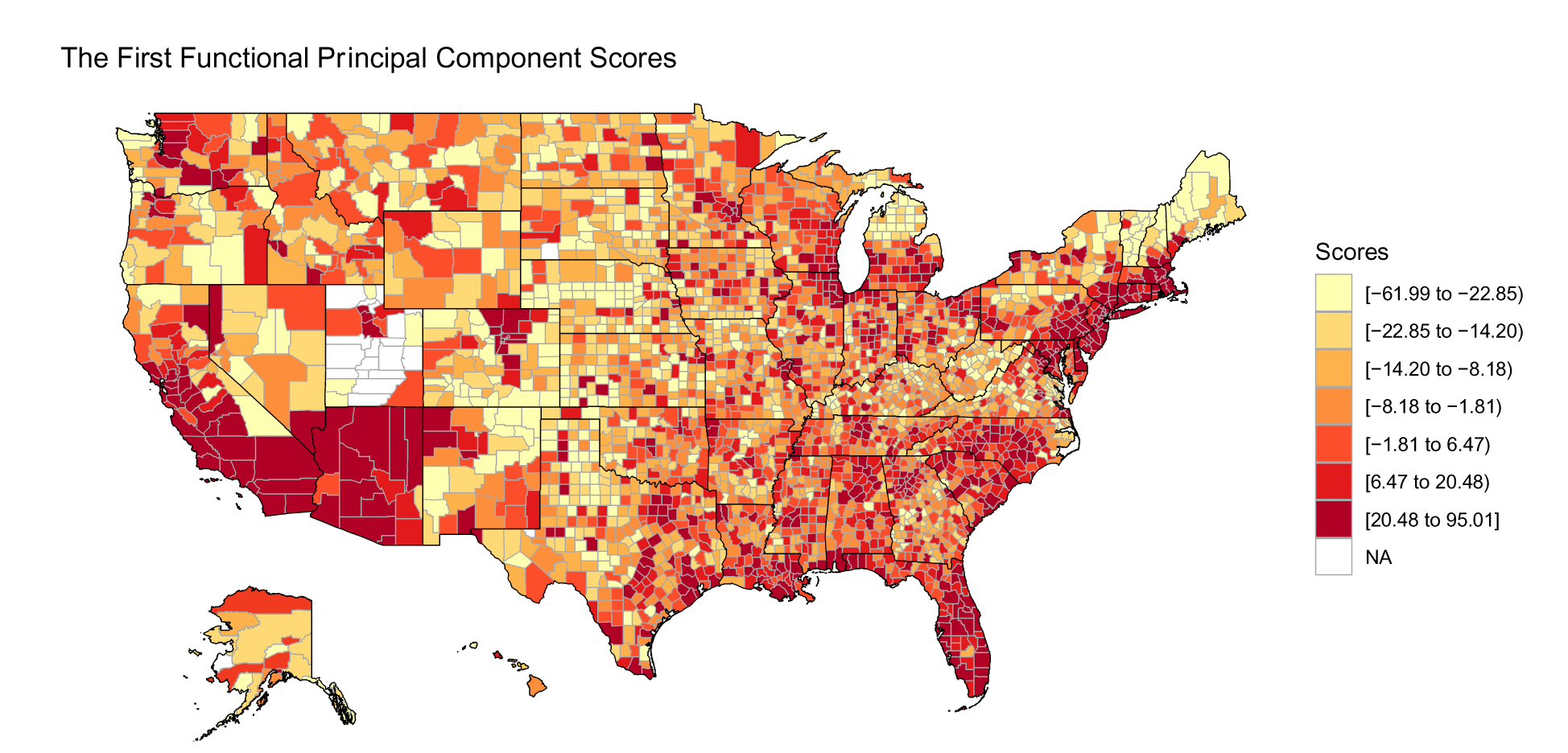
Heat map of the United States (US) according to the first FPC scores of counties.

### The Marginal Effects of Risk Factors

To identify the important features of the counties that influenced the pattern of COVID-19 trajectories over time, we determined the relationship between the first FPC score and the various characteristics of these counties using linear regression models. Table 2 summarizes the results of the linear regression models, which include regression coefficients, p-values, and the R^2^ Statistics. Among the 23 factors we investigated, all of these except ‘Proportion of Pacific Natives’ demonstrated a significant coefficient (p-value <0.005) for the first FPC score. The marginal R2 was moderate for these factors, up to 0.36. The variable ‘Number of Families’(R^2^=0.356) displayed the strongest association, which was followed by the variable, ‘Aggregate Family Income’ (R^2^=0.336). The two most negatively correlated factors were ‘Median Age’ (R^2^= 0.183) and ‘Proportion of Whites’ (R^2^= 0.0974).

### The Effects of Implementing a Lockdown

Figure S2-S25 in supplementary documents are scatter plots that depict the relationship of FPCs with various characteristics of the countries. Here, we observed a strong non-linear trend in the relationship between the first FPC scores and the timing of the lockdown. To better characterize this relationship, we used a segmented regression model. Compared with a linear regression model, segmented regression improved the fit of the model (i.e., *R*^2^**=**0.45 for segmented regression vs. *R*^2^=0.20 for linear regression). To evaluate the impact of a lockdown for each of these counties, we created three new variables (‘*Lockdown Indicator’, ‘Lockdown Time before the Inflection Point’, and ‘Lockdown Time after the Inflection Point’*), which were derived from the lockdown information of each county. Figure 3 shows the relationship of lockdown time with the first FPC scores. The blue dots denote the relationship between the lockdown time and the first FPC scores for each of the counties that used lockdowns to control the spread of COVID-19 infection. The red lines show the segments of a fitted line, which appeared as a “hockey stick” with an inflection point. Using time zero as the date on which a county reported at least 5 total cases of COVID-19, we identified day - 7.70 (i.e., approximately a week before a county reported 5 cumulative cases of COVID-19) as the average “inflection” point (the green vertical line in Figure 3) in the segmented regression model. The detailed results of the segmented regression model are shown in Table 2. Specifically, the coefficients of the variables, *‘Lockdown Slope before the Inflection Point’ and ‘Lockdown Slope after the Inflection Point’* in the segmented regression were all positive, corresponding to 0.0524, and 1.950, respectively.

**Figure 3:**
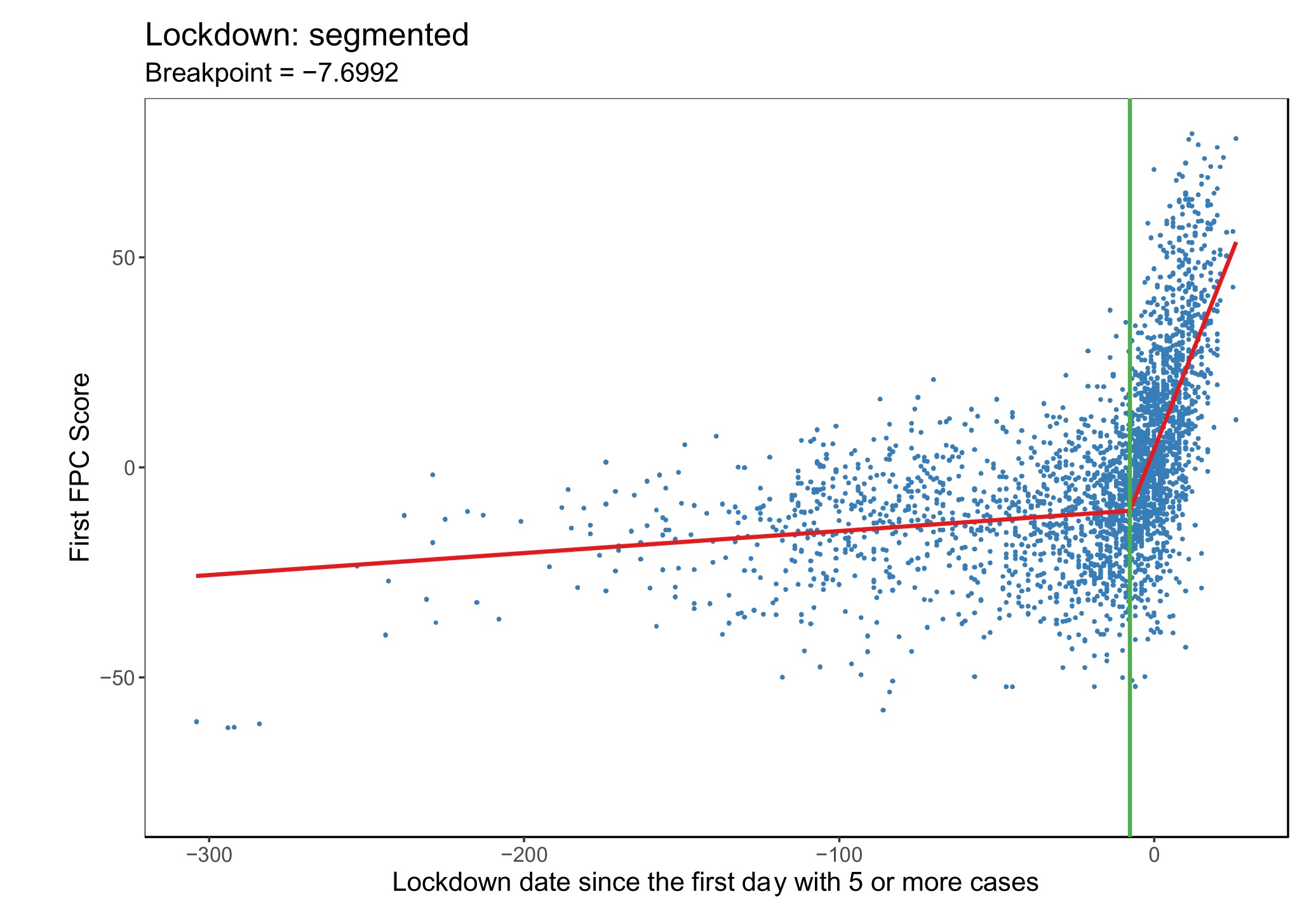
Relationship between the first FPC score and the lockdown date. The x-axis represents the number of days between the lockdown date and the date on which the county reported at least 5 COVID-19 cases. Positive values denote counties that instituted a lockdown after they reported at least 5 cumulative COVID-19 cases, while negative values denote counties that instituted a lockdown before they reported at least 5 cumulative COVID-19 cases. Each blue point represents data of a US county. The red lines represent two fitted slopes of a segmented regression model. The vertical green line (at −7.7 days) indicates the inflection point on which the slope of the first FPC score significantly changes.

### Joint modelling for all risk factors for COVID-19

We found that certain risk factors were highly correlated with each other as shown in the supplementary Figure S1. As seen in regression models, most of the risk factors we investigated were significantly associated with COVID-19 infection. To investigate their joint effects after adjusting for all of the other variables in Table 2, we used an elastic net model to determine the relationship of the first FPC scores with these predictors. The confidence intervals, obtained from 1000 bootstraps, are shown in Figure 4 and the mean value and 95% confidence interval of the optimal model’s coefficients are shown in supplementary Table S2. We note that the elastic net models, which incorporate the joint effects of different risk factors, achieved a much better *R*^2^ of 0.63, compared with the marginal regression results in which the maximal *R*^2^ was 0.36 for the first FPC score.

**Figure 4:**
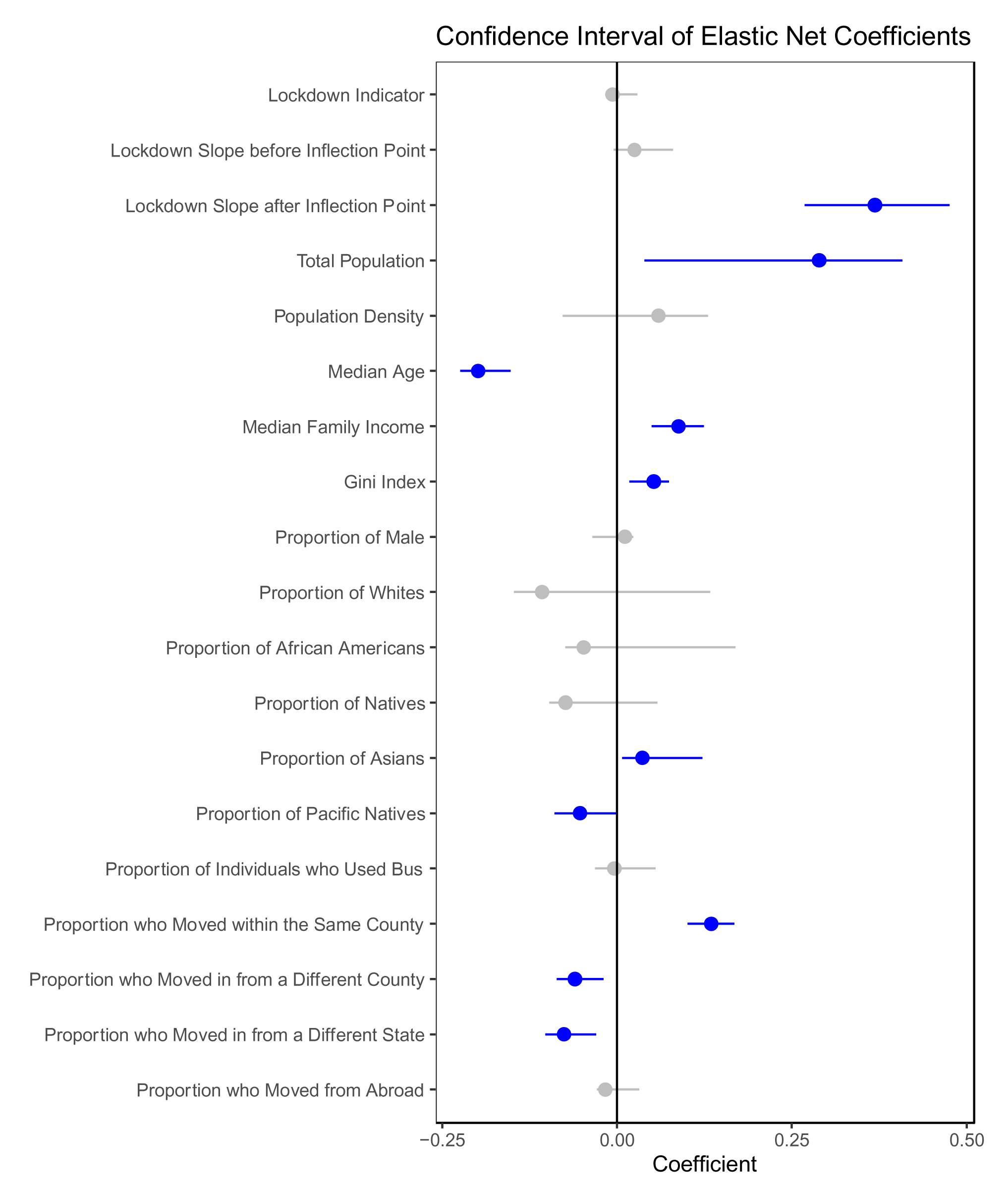
The adjusted relationship between characteristics of counties and the first FPC scores, based on results of elastic net models. The effect of every variable is adjusted to other factors listed in the figure. A positive coefficient denotes variables that are positively related to the number of COVID cases. The dot indicates the mean coefficients, and the bar represents the 95% confidence interval. Blue color indicates the significant factors whose 95% confidence interval does not cover 0.

We observed that 10 of 19 risk factors demonstrated statistical significance (i.e., consistently different from zero). For example, ‘*Lockdown Time after the Inflection point’* and ‘*Total Population’* were positively associated with the first FPC scores, while ‘*Median Age’* was negatively associated with the first FPC scores. Other positive risk factors included ‘*Median Family Income’, ‘Gini Index’, ‘Proportion of Asians’, and ‘Proportion who Moved within the Same County’*, whereas the other negative risk factors were ‘*Proportion of Pacific Natives’, ‘Proportion who Moved from a Different County’, and ‘Proportion who Moved from a Different State’*. Many other features became statistically insignificant in the joint models.

In the elastic net models, the mean slope for initiation of a lockdown after inflection point was 0.369, which was obtained from 1000 bootstraps. This indicates that, after adjusting for other important factors, if a lockdown was implemented after the inflection point, delays in lockdown were associated with an exponential increase in the COVID-19 case counts in the community. For example, in the first 62 days after time zero, each day of delay in lockdown led to an increase in the county case count of 0.827% to 2.08%. After 62 days, the rate of increase became constant at 2.11%. Each week of delay in lockdown in the first 62 days was associated with an increase in the COVID-19 case count of 5.93% to 15.5%; after which the weekly increase became constant at 15.7%.

## DISCUSSION

Lockdowns are an effective way of reducing the reproduction number of COVID-19 and controlling the disease in local communities. However, there is no consensus on when governments should take this action. Here, we found that communities, which implemented the lockdown at or prior to the inflection point (in which 5 cases were ascertained) experienced a slower rise in COVID-19 rates over 1 year. Communities that issued a lockdown after this inflection point experienced an exponential growth in their COVID-19 rates for every day they delayed their decision for up to 2 months; after which the growth rate became constant. In our models, the timing of the lockdown at the county level explained 44% of the total variance in the cases counts of COVID-19 in those communities.

We also found that certain characteristics of counties also significantly modified the case rates of COVID-19. These included variables that captured the wealth and population size of counties. Densely populated counties with a high median family income experienced a faster growth of COVID-19 in their communities. Interestingly, counties with a lower median age and a predominance of whites had lower growth rate of COVID-19, while the effect of whites proportion become non-significant after adjust to other factors. Conversely, counties that had more Asians experienced a faster growth rate of COVID-19. These data are consistent with those from the Centre for Disease Control, which has shown that the COVID-19 mortality rates are 10% higher in Asian-Americans and nearly 30% lower in the Whites, after age adjustments^17^. These data are also consistent with the observation that case rates decrease with increasing age in adulthood until 80 years of age, when the rates once again increase^18^. This is in contrast to COVID-19 mortality, which increases with advancing age with nearly 50% of all deaths occurring in those 75 years of age or older^19^. We found that the use of public transportation and increased mobility within counties also increase COVID-19 growth rates. However, these factors, though statistically significant, had a relatively modest impact on the overall growth rates. By far the more important predictors were the timing of the lockdown and the total population in counties.

Our findings extend those of two recent studies^1,2^ that had investigated the relationship of COVID-19 spread in communities with their population characteristics. Both studies, however, were cross-sectional in design, which limited causal inference. Here, we used a longitudinal design to evaluate the effects of lockdown and its joint effects with other population characteristics. Using this approach, we showed that the 1^st^ eigen-function is not a constant and thus the rate of COVID-19 growth changes from day to day. Importantly, we showed that the average slope in the growth rate markedly increased at an “inflection” point, which occurred approximately 7 days before counties ascertained at least 5 COVID-19 in their communities. After this inflection point, there was an exponential increase in COVID-19 counts unless lockdowns were implemented aggressively and quickly.

Another novel feature of the present study was by using a FPC analysis, we were able to convert the trajectory of each county into the first FPC score, which accounted for 93% of the total variance in the COVID-19 infection growth trajectories among US counties. This enabled us to use the first FPC score as a surrogate for infection case counts in these counties and model the relationship of the longitudinal COVID-19 infection pattern with the timing of lockdowns and other population characteristics of US counties. Using this approach, we also showed the impact of population size of counties on their COVID-19 trajectories. The heat map reveals that the most populous states, such as New York, California, and Florida, were the most impacted by SARS-CoV2, the virus responsible for COVID-19. At a city level, Los Angeles had the highest first FPC score, followed by Chicago, Miami, and New York, all with dense populations.

There were limitations to the study. First, as this was not a randomized controlled trial, unmeasured confounders could have distorted the overall findings. To minimize this possibility, we evaluated only counties in the US and adjusted for the most important characteristics of these counties. Second, although we chose the date of lockdown as the day on which the local government issued a “stay-at-home” order, other dates such as the date of a travel ban or school closing could also have been chosen. In a sensitivity analysis, we considered several of these possibilities (e.g. school closing, workplace closing, cancellation of public events, restrictions on gatherings, etc.) and found that the use of these dates produced very similar results to those of our main findings. Third, these data were generated in the US and may not apply to other countries around the world, which may have different characteristics and attitudes and adherence to public health policies such as masking and social distancing. Fourth, we did not consider the adherence of the residents in the counties to the lockdown order and did not evaluate the possible adverse effects of lockdowns including loss of income and productivity, mental health issues and “lockdown fatigue”.

Notwithstanding these limitations, our findings have important public health implications. Local state and municipal governments should issue an immediate lockdown order when only a few cases of COVID-19 are ascertained in their communities (less than 5); any significant delays in lockdown beyond this point are associated with an exponential increase in COVID-19 spread. The maximal rate of COVID-19 spread is, on average, achieved approximately 2 months following this inflection point.

## Data Availability

Data used in the manuscript are from online resource accessible to public

## Authors and Contributions

Xiaojian Shao, Li Xing, Don D. Sin, Xuekui Zhang contributed to the study concept and design. Xiaolin Huang, Xiaojian Shao, Yushan Hu contributed to the acquisition of the datasets from online resources, data processing and data analysis. Xiaolin Huang wrote the first draft. All authors have developed drafts of the manuscript, approved the final draft of the manuscript, and meet the criteria for authorship as recommended by the International Committee of Medical Journal Editors. Don Sin and Xuekui Zhang supervised this project.

